# Antihypertensive Medication Adherence and Medical Costs, Health Care Utilization, and Labor Productivity among Persons with Hypertension

**DOI:** 10.1101/2024.04.15.24305866

**Authors:** Jun Soo Lee, Raul Segura Escano, Nicole L. Therrien, Ashutosh Kumar, Ami Bhatt, Lisa M. Pollack, Sandra L. Jackson, Feijun Luo

## Abstract

**Background:** Hypertension affects nearly half of U.S. adults, yet remains inadequately controlled in over three-quarters of these cases. This study aims to assess the association between adherence to antihypertensive medications and total medical costs, health care utilization, and productivity-related outcomes.

**Methods:** We conducted a retrospective cohort study using MarketScan databases, which included individuals aged 18–64 with non-capitated health insurance plans in 2019. Adherence was defined as ≥80% Medication Possession Ratio (MPR) for prescribed antihypertensive medications. We used a generalized linear model to estimate total medical costs, a negative binomial model to estimate health care utilization (emergency department visits and inpatient admissions), an exponential hurdle model to estimate productivity-related outcomes (number of sick absences, short-term disability [STD], long-term disability [LTD]), and a two-part model to estimate productivity-related costs in 2019 U.S. dollars. All models were adjusted for age, sex, urbanicity, census region, and comorbidities. We reported average marginal effects for outcomes related to antihypertensive medication adherence.

**Results:** Among 379,503 individuals with hypertension in 2019, 54.4% adhered to antihypertensives. Per-person, antihypertensive medication adherence was associated with $1,441 lower total medical costs, $11 lower sick absence costs, $291 lower STD costs, and $69 lower LTD costs. Per 1,000 individuals, medication adherence was associated with lower healthcare utilization, including 200 fewer ED visits and 90 fewer inpatient admissions, and productivity-related outcomes, including 20 fewer sick absence days and 442 fewer STD days.

**Conclusions:** Adherence to antihypertensives was consistently associated with lower total medical costs, reduced healthcare utilization, and improved productivity-related outcomes.

## INTRODUCTION

Hypertension elevates the risk for heart disease and stroke, two leading causes of death for people in the U.S.^1^ Almost half of U.S. adults have hypertension, and over three-quarters of these individuals do not have their blood pressure controlled to <130/80 mmHg.^2^ Prescription medication is recommended, along with lifestyle modifications, for nearly 80% of those with hypertension to achieve blood pressure control.^2^ Adherence to antihypertensive medications, i.e. consistent use as prescribed, plays a critical role in hypertension control.^3–6^ Medication adherence is multifactorial and may be influenced by socioeconomic and demographic, health care system, therapeutic, and patient-related factors.^4^ Improving adherence to antihypertensive medications among people with hypertension is crucial for improving national hypertension control rates.^5,6^

In addition to improved hypertension control, adherence to antihypertensive medications is associated with reduced risk of stroke, fewer hospitalizations, and decreased cardiovascular morbidity and mortality.^7–14^ Despite its importance, rates of adherence to antihypertensive medications are suboptimal, though estimates vary by method of measurement and population characteristics.^4^ Chang et al. 2015^15^ found that nearly two-thirds of U.S. adults with hypertension were considered adherent to their antihypertensive medications, with lower adherence rates observed among Medicare beneficiaries, women, and younger adults. Improving medication adherence may impact economic outcomes, including total medical costs,^16–18^ hospitalization and emergency department (ED) visits,^11,19–21^ and other indirect outcomes such as work productivity^22,23^ and mortality rates.^24,25^ Lost work productivity, attributed to chronic conditions and resulting in absenteeism or disability leave, imposes an economic burden on the patient, employers, and healthcare systems.^26,27^ Enhanced medication adherence among employees with hypertension may lead to fewer days of absence and health care utilization savings.^27–29^ Although some studies have documented costs and health care utilization associated with adherence to antihypertensive medications,^11,16–19^ none have investigated productivity-related outcomes– indirect benefits–associated with such adherence.

This study aims to investigate the economic burden associated with adherence to antihypertensive medications, including total medical costs, health care utilization, and labor productivity. Administrative commercial claims linked with employer-provided payroll databases from 2019 were utilized to examine both medical costs and productivity-related costs associated with medication adherence.

## METHODS

### Data

We used the 2019 MarketScan Commercial Claims and Encounters (CCAE) and Health and Productivity Management (HPM) databases.^30^ The CCAE database contains claims for inpatient, emergency department (ED), outpatient, and prescription drugs from enrollees and their dependents covered by employer-sponsored commercial health insurance plans. This database aggregates data from over 300 employers, more than 30 health plans, and over 500 hospitals across the U.S. The HPM database is derived from a subset of employers’ payroll database, including information on recreational, sick, and other absences, as well as absences related to short-term disability (STD) and long-term disability (LTD). Linking the HPM database to commercial claims information in the CCAE database enables researchers to analyze associations between medical conditions, medication adherence, total medical costs, health care utilization, and labor productivity outcomes. This study involved secondary data analysis using de-identified information and was categorized as non-research and thus exempt from Institutional Review Board review.

### Identification of individuals with hypertension

We included individuals aged 18–64 years, continuously enrolled in the 2019 MarketScan CCAE and HPM databases. Within this group, individuals with hypertension were defined if they had at least one hypertension diagnosis (International Classification of Diseases, Tenth Revision, Clinical Modification [ICD-10-CM]=I10–I15) and at least one antihypertensive drug claim in 2019 (Appendix Table 1). We excluded individuals with a history of pregnancy (Appendix Table 2) and/or those covered under capitated insurance.

**Table 1.** Summary statistics of individuals with hypertension by medication adherence status, MarketScan® Commercial Insurance, 2019^a^.

|  | All | Medication Adherence<sup>b</sup> | Medication Non-adherence | P-value |
| --- | --- | --- | --- | --- |
|  | N=379,503 (100%) | N=206,310 (54.4%) | N=173,193 (45.6%) |  |
| Age, mean (SD) | 51.1 (8.4) | 52.5 (7.7) | 49.4 (8.9) | <0.001 |
| **Age groups, n (%)** |  |  |  |  |
| 18-34 | 18,480 (4.87%) | 5,808 (2.82%) | 12,672 (7.32%) | <0.001 |
| 35-44 | 61,732 (16.27%) | 26,358 (12.78%) | 35,374 (20.42%) | <0.001 |
| 45-54 | 142,361 (37.51%) | 76,387 (37.03%) | 65,974 (38.09%) | <0.001 |
| 55-64 | 156,930 (41.35%) | 97,757 (47.38%) | 59,173 (34.17%) | <0.001 |
| Female, n (%) | 128,245 (33.79%) | 64,679 (31.35%) | 63,566 (36.70%) | <0.001 |
| Urban Residency, n (%) | 341,292 (89.93%) | 184,978 (89.66%) | 156,314 (90.25%) | <0.001 |
| **Census Region, n (%)** |  |  |  |  |
| Northeast | 52,646 (13.87%) | 31,090 (15.07%) | 21,556 (12.45%) | <0.001 |
| Midwest | 96,314 (25.38%) | 54,545 (26.44%) | 41,769 (24.12%) | <0.001 |
| South | 182,220 (48.02%) | 94,087 (45.60%) | 88,133 (50.89%) | <0.001 |
| West | 47,611 (12.55%) | 26,286 (12.74%) | 21,325 (12.31%) | <0.001 |
| **Comorbidities, n (%)** |  |  |  |  |
| Myocardial infarction | 6,951 (1.83%) | 3,468 (1.68%) | 3,483 (2.01%) | <0.001 |
| Congestive heart failure | 7,549 (1.99%) | 3,547 (1.72%) | 4,002 (2.31%) | <0.001 |
| Peripheral vascular disease | 5,493 (1.45%) | 2,980 (1.44%) | 2,513 (1.45%) | 0.87 |
| Cerebrovascular disease | 1,775 (0.47%) | 749 (0.36%) | 1,026 (0.59%) | <0.001 |
| Dementia | 49 (0.01%) | 19 (0.01%) | 30 (0.02%) | 0.028 |
| Chronic pulmonary disease | 16,189 (4.27%) | 8,117 (3.93%) | 8,072 (4.66%) | <0.001 |
| Rheumatic disease | 3,981 (1.05%) | 2,116 (1.03%) | 1,865 (1.08%) | 0.12 |
| Peptic ulcer disease | 719 (0.19%) | 324 (0.16%) | 395 (0.23%) | <0.001 |
| Mild liver disease | 6,196 (1.63%) | 3,079 (1.49%) | 3,117 (1.80%) | <0.001 |
| Diabetes without chronic complication | 69,362 (18.28%) | 40,754 (19.75%) | 28,608 (16.52%) | <0.001 |
| Diabetes with chronic complication | 15,073 (3.97%) | 8,584 (4.16%) | 6,489 (3.75%) | <0.001 |
| Hemiplegia or paraplegia | 802 (0.21%) | 270 (0.13%) | 532 (0.31%) | <0.001 |
| Renal disease | 9,818 (2.59%) | 5,210 (2.53%) | 4,608 (2.66%) | 0.009 |
| Any malignancy | 10,373 (2.73%) | 5,972 (2.89%) | 4,401 (2.54%) | <0.001 |
| Moderate or severe liver disease | 468 (0.12%) | 182 (0.09%) | 286 (0.17%) | <0.001 |
| Metastatic solid tumor | 1,379 (0.36%) | 657 (0.32%) | 722 (0.42%) | <0.001 |
| AIDS/HIV | 1,256 (0.33%) | 694 (0.34%) | 562 (0.32%) | 0.53 |
| **Outcomes, mean (SD), \$** | | | | |
| Total medical costs | 14081.8 (40871.8) | 12882.2 (32497.4) | 15510.7 (48976.3) | <0.001 |
| Total insurance medical payments | 12425.0 (40176.8) | 11250.1 (31776.9) | 13824.5 (48275.8) | <0.001 |
| Total individuals' out-of-pocket medical costs | 1656.8 (1932.0) | 1632.1 (1756.2) | 1686.3 (2122.2) | <0.001 |
| Sick absence costs | 76.6 (807.9) | 70.6 (754.9) | 83.9 (866.9) | <0.001 |
| STD costs | 686.5 (3676.0) | 533.8 (3143.7) | 868.5 (4216.3) | <0.001 |
| LTD costs | 159.9 (4232.6) | 120.3 (2766.5) | 207.1 (5489.5) | <0.001 |
| Num. of emergency department visits | 0.34 (0.99) | 0.25 (0.78) | 0.46 (1.18) | <0.001 |
| Num. of inpatient admissions | 0.09 (0.41) | 0.06 (0.32) | 0.12 (0.49) | <0.001 |
| Num. of outpatient visits | 12.90 (16.05) | 12.73 (15.29) | 13.11 (16.90) | <0.001 |
| Num. of pharmacy prescriptions | 25.21 (20.05) | 28.30 (20.78) | 21.53 (18.49) | <0.001 |
| Num. of sick absences | 2.38 (9.25) | 2.22 (8.70) | 2.57 (9.85) | <0.001 |
| Num. of STD | 5.07 (25.15) | 3.93 (21.49) | 6.43 (28.88) | <0.001 |
| Num. of LTD | 1.40 (34.12) | 1.06 (22.36) | 1.80 (44.12) | <0.001 |
| Num. of antihypertensive prescriptions | 7.93 (6.42) | 9.43 (6.80) | 6.14 (5.42) | <0.001 |

Abbreviations: ED, emergency department; LTD, long-term disability; STD, short-term disability.
<sup>a</sup> The Wilcoxon nonparametric rank-sum test was used to test the differences in means for continuous variables, and the Pearson's Chi-square test was used to test differences in proportions for categorical variables by medication non-adherence to antihypertensives status.
<sup>b</sup> Adherence to antihypertensives if the average medication possession ratio (MPR) of the 7 Therapeutic Classes was greater than equal to 80%; otherwise, medication non-adherence.

**Table 2.** Total medical costs (per individual) associated with adherence to antihypertensives in 2019^a^.

|  | All | Women | Men | Women vs. Men<sup>b</sup> | Urban | Rural | Urban vs. Rural<sup>b</sup> |
| --- | --- | --- | --- | --- | --- | --- | --- |
| **Total Medical Costs, \$** | | | | | | | |
| Medication non-adherence | 19,210<br>(18,643 - 19,778) | 20,488<br>(19,870 - 21,107) | 18,558<br>(17,957 - 19,160) | 1,930***<br>(1,474 - 2,386) | 19,498<br>(18,907 - 20,089) | 16,641<br>(15,977 - 17,306) | 2,857***<br>(2,221 - 3,493) |
| Medication adherence | 17,770<br>(17,238 - 18,301) | 18,837<br>(18,265 - 19,408) | 17,225<br>(16,663 - 17,787) | 1,611***<br>(1,205 - 2,017) | 17,971<br>(17,419 - 18,523) | 15,970<br>(15,373 - 16,566) | 2,002***<br>(1,454 - 2,550) |
| Difference | -1,441***<br>(-1,709 - -1,172) | -1,652***<br>(-2,131 - -1,172) | -1,333***<br>(-1,650 - -1,016) | -319<br>(-886 - 249) | -1,527***<br>(-1,813 - -1,241) | -671.7<br>(-1,399 - 55.76) | -855*<br>(-1,633 - -77) |
| Observation | 379,503 | 128,245 | 251,258 | 379,503 | 341,292 | 38,211 | 379,503 |

<sup>a</sup> A generalized linear model with a gamma distribution and log link was used. The average marginal effects with 95% confidence intervals (CI) were reported. All models were adjusted for the patient's age, sex, urbanicity of residence, Census regions, and comorbidities. The differences and the 95% CI of the predicted and average marginal effects by sex and urbanicity were calculated. All the outcomes are per individual.
<sup>b</sup> The differences with 95% CI in the predicted values and average marginal effects by sex (Women – Men) and urbanicity (Urban – Rural) were reported.
\*\*\* p<0.001, \*\* p<0.01, \* p<0.05

### Identification of adherence to antihypertensive medications

Antihypertensive medications (antihypertensives) were identified using generic names and categorized by therapeutic class (ACE inhibitors, angiotensin receptor blockers, beta-blockers, calcium channel blockers, diuretics, other antihypertensives, and renin-angiotensin system antagonists [ACE inhibitors, angiotensin receptor blockers, and direct renin inhibitors]) (Appendix Table 1). The average medication possession ratio (MPR) for antihypertensives by therapeutic class in 2019 was calculated as the ratio of the sum of the days’ supply for all drug fills to the number of days in the study period.^31^ For individuals taking multiple antihypertensives, the MPR was calculated as an average of the MPR for each therapeutic class. Adherence was defined as an average MPR greater than or equal to 80%, while non-adherence was indicated by an average MPR below 80%.^31^ A dummy indicator of adherence to antihypertensives was created, with a value of one for adherent individuals and zero for non-adherent individuals.

### Outcome variables

Dependent variables included 1) total all-cause medical costs (the sum of individuals’ out-of-pocket costs and insurance payments), 2) health care utilization measured by the number of ED visits and inpatient admissions, and 3) productivity-related outcomes quantified by the number of sick absences and absences related to STD and LTD. Our focus was on sick absences rather than other types, such as recreational, jury duty, military leave, or plant shutdowns, which may not be directly affected by medication adherence. Productivity-related costs were calculated by multiplying absence hours by average hourly wages in 2019^32^, with adjustments: 100% for absences, 70% for STD-related absences, and 60% for LTD-related absences.^32,33^ Average hourly wages in 2019 U.S. dollars ($27.99) were sourced from the U.S. Bureau of Labor Statistics (BLS) for all employees on private nonfarm payrolls.

### Statistical analysis

We categorized individuals into age groups (18-34 [reference], 35-44, 45-54, and 55-64), sex (male [reference] and female), urbanicity (rural [reference] and urban), census regions (Northeast [reference], Midwest, South, and West), and comorbidities, specifically the Quan 17 Charlson Comorbidities.^34^ Comorbidities were defined if individuals had at least one inpatient admission or two outpatient visits with a 30-day interval.

Summary statistics were documented for age, sex, urbanicity and census region of residence, and comorbidities. Differences in average values by medication adherence status were tested using the Wilcoxon rank-sum test for continuous variables and Pearson’s Chi-square test for categorical variables.

For total medical costs, a generalized linear model with a log link and gamma distribution was used. For costs associated with productivity-related outcomes, we utilized a two-part model. The first part was a logit model, and the second part was a generalized linear model with a log link and gamma distribution. For health care utilization (number of ED visits and inpatient admissions), a negative binomial model was used. For productivity-related outcomes, where they contained excess zeros, we employed an exponential hurdle model. All models were adjusted for individuals’ age, sex, urbanicity, census regions, and dummy indicators for the 17 Charlson comorbidities. Average marginal effects of outcomes associated with adherence to antihypertensives were reported.

For the sensitivity analysis, we employed the overlap weighting method so measured confounders were equally distributed between the adherence and non-adherence groups.^35–38^ The overlap weighting method mimics random assignment by creating a pseudo-population through weighting so that measured confounders are equally distributed between adherence and non-adherence groups.^36,37^ We calculated overlap weights that were proportional to the probability of individuals belonging to the opposite group. Weights were obtained from propensity scores, estimated by logistic regression with medication adherence as the outcome. We used the iterative method to estimate propensity scores proposed by Imbens and Rubin.^39,40^ This method involved utilizing baseline covariates, as reported in the main model, to perform higher-order interactions for covariate selection in logistic regression, maximizing the likelihood function. Estimates of logistic regression, assessment of covariate balance, and probability distributions for weighted samples for overall and by sex, age group and urbanicity are presented in Appendix Tables 4-6 and Appendix Figures 1-4. *P*-values less than 0.05 were considered statistically significant. All analyses were conducted by using Stata SE version 17 (StataCorp, College Station, TX) in 2023.

**Figure 1.**
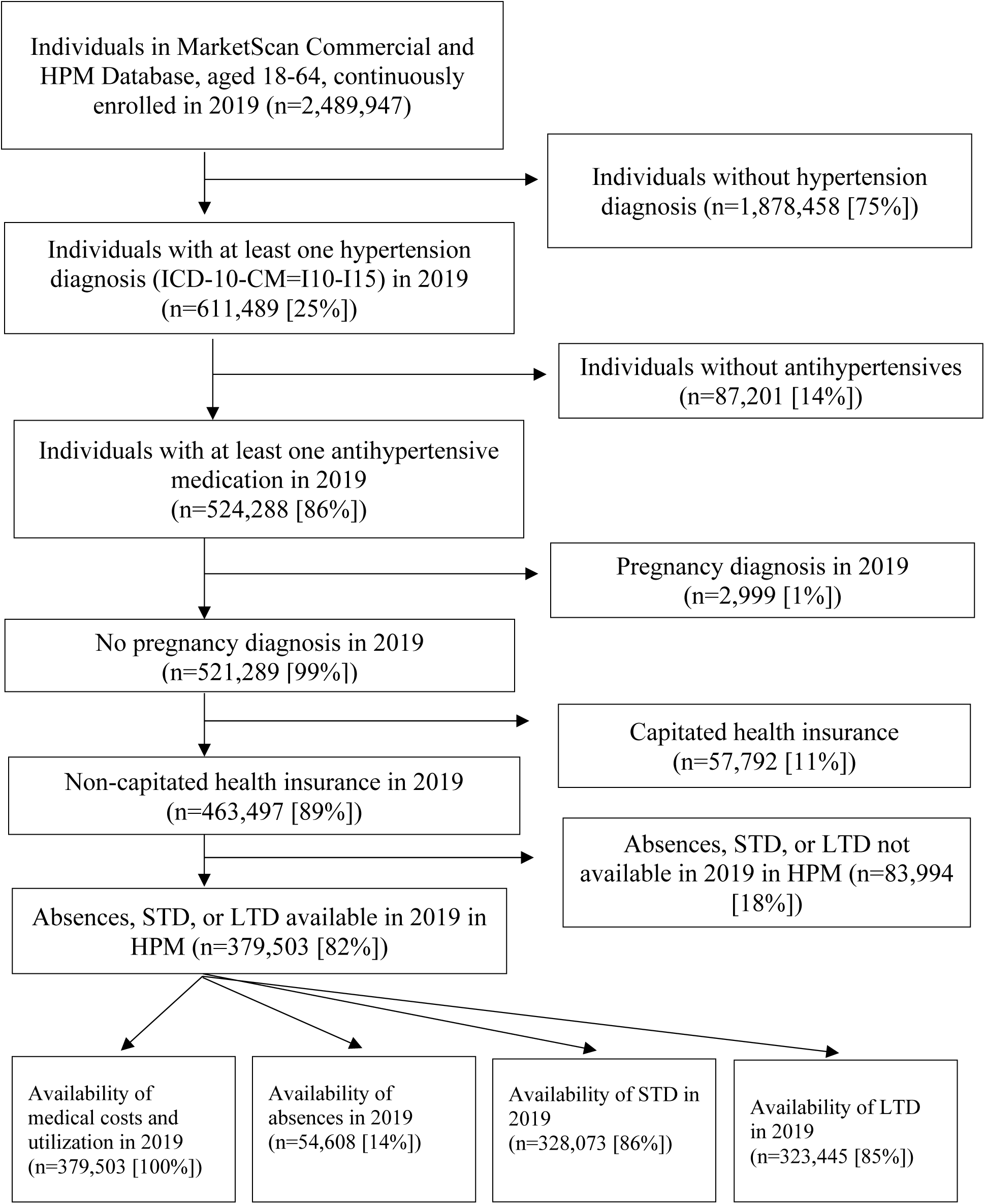
Study sample selection of individuals diagnosed with hypertension, MarketScan® Commercial and Health and Product Management Database, 2019.

## RESULTS

A total of 379,503 individuals with hypertension were identified who had availability of absence, STD, or LTD information (Figure 1). All 379,503 individuals had information on medical costs, health care utilization, and drug claims. Among these individuals, 54,608 (14%) had absence information, 328,073 (86%) had STD information, and 323,445 (85%) had LTD information.

Out of the 379,503 individuals, 206,310 (54.4%) individuals adhered to antihypertensives (Table 1). Those who adhered (mean [SD] age, 52.5 [7.7] years) were older than those who did not (mean [SD] age, 49.4 [8.9]; *P*<0.001). Adherent individuals were less likely to be female (31.35% vs. 36.70%; *P*<0.001). Medication adherence rates were also lower for females than males (50.43% vs 56.37%; *P*<0.001), and for individuals living in urban areas compared to those in rural areas (54.20% vs 55.83%; *P*<0.001) (Appendix Table 7). Adjusted results showed that females were 5.83% (95% CI, 6.16 to 5.50; *P*<0.001) less likely to adhere to medications than males, and urban residents were 1.38% (95% CI, −1.89 to −0.86; *P*<0.001) less likely to adhere to medications than those residing in rural areas (Appendix Table 8).

Table 2 presents the total medical costs associated with adherence to antihypertensives. The predicted total medical cost for individuals who adhered to antihypertensives was $17,770 (95% CI, 17,238 to 18,301), while for those who did not adhere, the predicted total medical cost was $19,210 (95% CI, 18,643 to 19,778). Thus, compared to individuals who did not adhere, those who adhered had a $1,441 per individual lower total medical costs (95% CI, −1,709 to −1,172; *P*<0.001). The association of medication adherence with total medical costs was more pronounced for individuals residing in urban areas, showing an additional decrease of $855 (95% CI, −1,633 to −77; *P*<0.05), compared with those living in rural areas.

Table 3 documents the number of ED visits and inpatient admissions per 1,000 individuals associated with adherence, both overall and stratified by sex and urbanicity of residence. Adherence to antihypertensives was associated with 200 fewer ED visits per 1,000 individuals (95% CI, −206 to −194; *P*<0.001) and 89.6 fewer inpatient admissions per 1,000 individuals (95% CI, −95.5 to −83.7; *P*<0.001). Women experienced an additional 113 fewer ED visits (95% CI, −126 to −99.5; *P*<0.001) associated with medication adherence compared to men, while men had an additional 26.6 fewer inpatient admissions (95% CI, 17.9 to 25.3; *P*<0.001) associated with medication adherence compared to women. Individuals living in urban areas had fewer inpatient admissions associated with medication adherence than those living in rural areas by 35.5 (95% CI, −46.8 to −24.2; *P*<0.001).

**Table 3.**
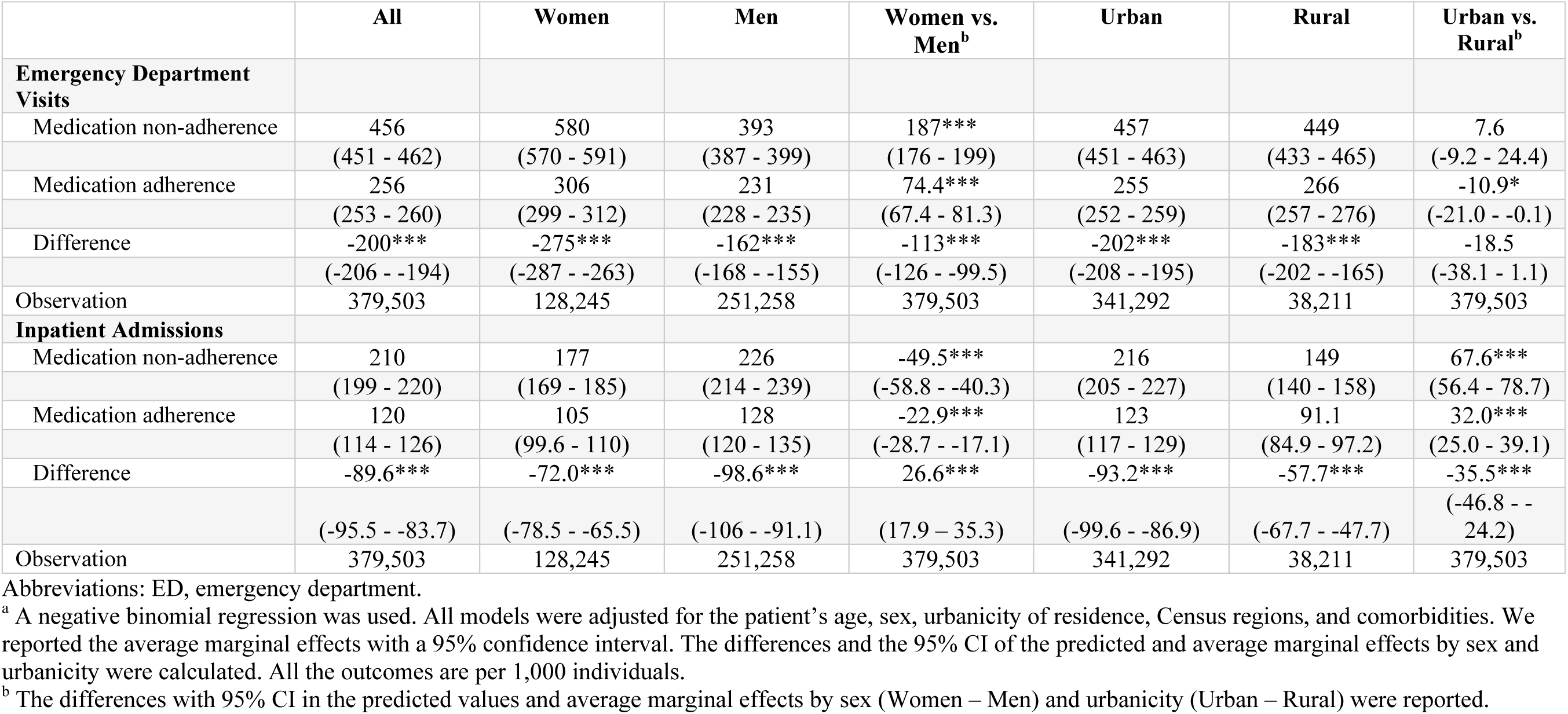
Health care utilization (per 1,000 individuals) associated with adherence to antihypertensives in 2019^a^.

Table 4 presents productivity losses per 1,000 individuals associated with medication adherence. Individuals adhering to antihypertensives had fewer sick absences (−19.8 days per 1,000 individuals, 95% CI, −28.0 to −11.6; *P*<0.001) and STD days (−442 days, 95% CI, −548 to −335; *P*<0.001). The association between medication adherence and the number of sick absences was more pronounced for individuals living in rural areas, showing a stronger association in sick absences by 46.6 days (95% CI, 11.3 to 81.9; *P*<0.001) than those living in urban areas.

**Table 4.** Productivity losses (per 1,000 individuals) associated with adherence to antihypertensives in 2019^a^.

|  | All | Women | Men | Women vs. Men<sup>b</sup> | Urban | Rural | Urban vs. Rural<sup>b</sup> |
| --- | --- | --- | --- | --- | --- | --- | --- |
| **Number of Sick Absences** |  |  |  |  |  |  |  |
| Medication non-adherence | 308 | 329 | 297 | -31.6*** | 299 | 385 | -86.0*** |
|  | (300 - 316) | (316 - 341) | (288 - 306) | (-44.8 - -18.3) | (291 - 307) | (357 - 413) | (-114 - -57.9) |
| Medication adherence | 288 | 306 | 279 | -26.8*** | 284 | 323 | -39.4*** |
|  | (281 - 295) | (294 - 318) | (271 - 286) | (-39.2 - -14.4) | (277 - 291) | (302 - 345) | (-61 - -17.8) |
| Difference | -19.8*** | -22.9** | -18.2*** | 4.75 | -15.1*** | -61.7*** | 46.6** |
|  | (-28.0 - -11.6) | (-38.7 - -7.2) | (-27.4 - -8.9) | (-13.3 - 22.8) | (-23.3 - -6.9) | (-96.1 - -27.3) | (11.3 - 81.9) |
| Observation | 54,608 | 13,346 | 41,262 | 54,608 | 49,125 | 5,483 | 54,608 |
| **Number of STD** |  |  |  |  |  |  |  |
| Medication non-adherence | 4,514 | 4,354 | 4,596 | 241** | 4,548 | 4,211 | 337** |
|  | (4,414 - 4,615) | (4,230 - 4,479) | (4,475 - 4,716) | (96.7 - 386) | (4,444 - 4,652) | (3,976 - 4,446) | (96.8 - 577) |
| Medication adherence | 4,072 | 3,925 | 4,147 | 222** | 4,086 | 3,950 | 137 |
|  | (3,978 - 4,166) | (3,799 - 4,052) | (4,036 - 4,259) | (75 - 369) | (3,988 - 4,184) | (3,731 - 4,168) | (-88.7 - 362) |
| Difference | -442*** | -429*** | -448*** | -19.2 | -462*** | -262 | -200 |
|  | (-548 - -335) | (-585 - -273) | (-586 - -311) | (-224 - 186) | (-575 - -349) | (-572 - 48.5) | (-529 - 128) |
| Observation | 328,073 | 111,160 | 216,913 | 328,073 | 295,104 | 32,969 | 328,073 |
| **Number of LTD** |  |  |  |  |  |  |  |
| Medication non-adherence | 1,508 | 1,233 | 1,648 | 415*** | 1,479 | 1,761 | -281 |
|  | (1,319 - 1,697) | (1,045 - 1,421) | (1,412 - 1,884) | (173 - 657) | (1,289 - 1,669) | (1,243 - 2,278) | (-794 - 232) |
| Medication adherence | 1,624 | 1,445 | 1,715 | 269 | 1,596 | 1,870 | -274 |
|  | (1,414 - 1,833) | (1,187 - 1,704) | (1,460 - 1,970) | (-43.4 - 582) | (1,382 - 1,810) | (1,314 - 2,427) | (-836 - 287) |
| Difference | 116 | 212 | 66.8 | -146 | 117 | 110 | 6.8 |
|  | (-94.2 - 326) | (-64.2 - 489) | (-215 - 348) | (-537 - 245) | (-102 - 335) | (-618 - 838) | (-752 - 766) |
| Observation | 323,445 | 110,894 | 212,551 | 323,445 | 290,853 | 32,592 | 323,445 |

Abbreviations: LTD, long-term disability; STD, short-term disability.
<sup>a</sup> An exponential hurdle model was used. All models were adjusted for the patient's age, sex, urbanicity of residence, Census regions, and comorbidities. We reported the average marginal effects with a 95% confidence interval. The differences and the 95% CI of the predicted and average marginal effects by sex and urbanicity were calculated. All the outcomes are per 1,000 individuals.
<sup>b</sup> The differences with 95% CI in the predicted values and average marginal effects by sex (Women – Men) and urbanicity (Urban – Rural) were reported.

Appendix Table 3 presents the productivity costs per individual associated with medication adherence. Individuals adhering to antihypertensives had $11 lower costs associated with sick absences (95% CI, −16 to −6; *P*<0.001), $291 lower costs associated with STD (95% CI, −315 to −268; *P*<0.001), and $69 lower costs associated with LTD (95% CI, −97 to −41; *P*<0.001) compared to those who did not adhere. Women experienced a stronger association between STD costs and medication adherence ($72, 95% CI, −121 to −22; *P*<0.01) than men, and individuals living in urban areas had a stronger association in costs associated with STD (-$173, 95% CI, −249 to −97; *P*<0.001) than those living in rural areas.

Sensitivity analyses using overlap-weighted model showed overall results similar to the main results (Appendix Tables 9-12). Specifically, compared with those who did not adhere, individuals adhering to antihypertensives had a $1,378 lower total medical costs per individual (95% CI, −1,628 to −1,128; *P*<0.001) and 82 fewer inpatient admissions per 1,000 individuals (95% CI, −87 to −77; *P*<0.001) (Appendix Tables 9 and 10). Appendix Tables 11 and 12 present productivity-related outcomes, demonstrating consistent results.

## DISCUSSION

Using administrative linked commercial claims and employer-provided payroll databases, we examined total medical costs, health care utilization, and labor productivity associated with adherence to antihypertensives. Individuals with higher adherence rates had significantly lower total medical costs ($1,441 per individual), along with fewer ED visits (200 fewer visits per 1,000 individuals per year) and fewer inpatient admissions (89.6 fewer admissions per 1,000 individuals per year) compared with their non-adherent counterparts. For productivity-related outcomes, individuals who adhered to antihypertensives had fewer sick absences, STD, and LTD, as well as lower related costs, compared with those who did not adhere. When analyzing the association of medication adherence among population subgroups, we observed differences by sex and urbanicity of residence.

Our findings align with existing literature on the association between medication adherence and reduced preventable health care utilization, decreased total medical costs, and improved productivity-related outcomes^22,23,29^. In a 2018 systematic review, Cutler et al.^16^ identified 12 studies focused on cardiovascular disease and total healthcare costs, revealing adjusted annual economic costs of non-adherence for cardiovascular disease ranged from $3,347 to $19,472 per person. Baker-Goering et al. in 2019^18^ found predicted expenditures to be $610 lower among individuals who were adherent to antihypertensives compared with those who were not adherent. While our findings indicate more modest savings, variations in the formula used to calculate medication adherence and the medication non-adherence cut-off points, as well as our focus on hypertension, likely contribute to these differences. With regard to productivity, Gifford et al. systematic review in 2018^41^ on adherence to antihypertensives demonstrated that employee populations with chronic conditions can generate productivity and health care utilization savings at the organization level as adherence increases. Our analysis aligns with these findings in productivity and health care utilization savings. However, differences in indicators, outcome measures, and chronic conditions may have influenced the magnitude of estimated savings.

Describing the economic costs of uncontrolled hypertension has been recognized as a strategic approach to elevate hypertension control to a national priority and improve hypertension control across the U.S.^5^ The association between medication adherence and total medical costs, health care utilization, and productivity-related outcomes can be explained through various potential mechanisms. Individuals who adhere to medications typically exhibit better control of chronic disease and health outcomes,^7–10,12–14,42^ leading to lower number of ED visits and inpatient admissions related to costly acute events, subsequently lowering total medical costs. Moreover, individuals with improved health outcomes tend to experience fewer sick absences^41^ and rely less on disability-related insurance, contributing to increased productivity and reduced costs to employers. However, it is crucial to acknowledge the potential influence of unmeasured social determinants of health (SDOH), especially economic factors such as poverty, unstable housing, food insecurity, lack of transportation, and lack of social support. These factors may have affected both lower medication adherence and medical costs, ED visits, sick absences, etc.^43–45^ While our study controlled for many confounding variables, the potential impact of these unmeasured SDOH indicators cannot be ignored and may represent an important area for future research.

Prior studies have highlighted variations in medication adherence based by sex.^46^ Women, while more likely to use medication annually than their male counterparts, were less likely to adhere to prescribed medication.^47^ This difference might be attributable to women more frequently experiencing side effects^48,49^ and managing complex regimens,^50^ despite their higher likelihood of seeking preventive care.^51^ A separate study found that, although men demonstrated higher rates of adherence than women in the age range of 20-40 years, this trend reversed later in life (50-70 years).^52^ Further research could help elucidate the reasons behind sex-based differences.

The Surgeon General’s Call to Action to Control Hypertension recognizes the prioritization of medication adherence as an evidence-based strategy to achieve improvements in hypertension control.^5^ Moreover, the Call to Action highlights several interventions designed to empower and equip patients for better medication adherence. These interventions include self-measured blood pressure monitoring, tailored interventions by pharmacists, shared decision-making and motivational interviewing, reducing or eliminating cost sharing for medications, simplifying medication regimens, including the use of fixed-dose combination antihypertensives, and synchronizing refills for multiple medications to a single date.^4,5,53^ Clinicians can implement or expand these interventions to support medication adherence in hypertension care.^4,5^ Employers can play a pivotal role by supporting coverage of these interventions in employer-sponsored health plans or implementing medication adherence support services in the workplace. Employers may find that the reductions in costs due to averted health care utilization and productivity losses may exceed program costs.^5,54^

This study has several limitations. First, the MarketScan CCAE database is not a random sample, and the results may not be fully generalizable to all individuals with commercial insurance. Second, our results excluded individuals without insurance or those publicly insured, such as Medicare and Medicaid beneficiaries. If individuals with public insurance or without insurance exhibit different patterns in medication adherence, our results may not be universally applicable. Additionally, our study was restricted to individuals aged 18-64, limiting generalizability to older (≥65) or younger (≤17) individuals. Third, our inclusion criteria required continuous enrollment, and individuals with gaps in insurance coverage or job disruptions during the period were not captured in our analysis. Fourth, while the MPR is a widely used method for measuring medication adherence, it may overestimate adherence for individuals who routinely refill medications early. MPR, based on drug claims data, captures prescription filling behavior but does not directly measure individuals’ medication-taking behavior. Fifth, our study focused exclusively on adherence to antihypertensives, and did not capture adherence to medications for other conditions that may impact health, and subsequent health care utilization and labor productivity. Sixth, although we conducted a sensitivity analysis using the propensity score overlap weighting method, with all results remaining consistent, due to the limited information available in the claims database, we were unable to include other potential confounding factors such as individuals’ education level, income, race, and poverty level. Seventh, the cross-sectional nature of our study may not capture longer-term effects of medication adherence. The observed higher associations on STD compared to LTD may be attributed to cross-sectional data; examining longitudinal data might reveal different associations with LTD. Seventh, STD costs were calculated using the same average hourly wages for working adults in the U.S. The decrease in the STD and LTD costs associated with medication adherence was notably higher for men and those living in urban areas. Given the higher hourly wages for men and urban residents, our estimates of productivity-related costs might be underestimated. As our estimates are derived from interactions of medication adherence, sex, and urban indicators, we could not account for different hourly wages by sex and urbanicity of residence.

Our study focused on outcomes available from administrative claims data and employer-provided payroll system; consequently, we could not look at individuals’ health outcomes. Future research endeavors could delve into the association of medication adherence with health outcomes, productivity-related outcomes, and other non-medical costs for individuals who are uninsured or publicly insured, extending the analysis over a longer time horizon. Additionally, our study focused solely on pre-COVID-19 pandemic periods, while future studies might broaden the scope to include pandemic periods when medication adherence may have been impacted by interruptions and delays in healthcare services.^55^

## CONCLUSIONS

Our study identified the associations of antihypertensive medication adherence with total medical costs, health care utilization, and labor productivity, including number of sick absences, STD, and LTD. Medication adherence was associated with fewer ED visits and inpatient admissions, lower total medical costs, and fewer sick absences, STD, and LTD. These findings highlighting not only the potential benefits of medication adherence in reducing medical costs but also improving productivity-related outcomes. The results emphasize the potential for implementing and expanding programs to support medication adherence among individuals with hypertension covered by commercial insurance.

## Data Availability

The data is not publicly available due to data user agreement, but the program codes are available upon request to the corresponding author.

## Supplemental materials

**Appendix Table 1.** Antihypertensives by therapeutic class.

| Therapeutic Class | Antihypertensive Medications |
| --- | --- |
| ACE inhibitor | Benazepril<br>Bepridil<br>Captopril<br>Enalapril<br>Fosinopril<br>Lisinopril<br>Moexipril<br>Perindopril<br>Quinapril<br>Ramipril<br>Trandolapril |
| Angiotensin receptor blocker | Azilsartan<br>Candesartan<br>Eprosartan<br>Irbesartan<br>Losartan<br>Olmesartan<br>Telmisartan<br>Valsartan |
| Beta blocker | Acebutolol<br>Atenolol<br>Betaxolol<br>Bisoprolol<br>Carvedilol<br>Labetalol<br>Metoprolol succinate<br>Metoprolol tartrate<br>Nadolol<br>Nebivolol<br>Pindolol<br>Propranolol |
| Calcium channel blocker | Amlodipine<br>Diltiazem<br>Felodipine<br>Isradipine<br>Levamlodipine<br>Nicardipine<br>Nifedipine<br>Nisoldipine<br>Verapamil |
| Diuretic | Amiloride<br>Bumetanide<br>Chlorothiazide |
|  | Chlorthalidone<br>Furosemide<br>Hydrochlorothiazide<br>Indapamide<br>Methyclothiazide<br>Metolazone<br>Torsemide<br>Triamterene |
| Other antihypertensives | Clonidine<br>Doxazosin<br>Eplerenone<br>Guanabenz<br>Guanfacine<br>Hydralazine<br>Methyldopa<br>Minoxidil<br>Prazosin<br>Spironolactone<br>Terazosin |
| Renin-angiotensin system antagonists* | Aliskiren<br>Azilsartan<br>Benazepril<br>Bepridil<br>Candesartan<br>Captopril<br>Enalapril<br>Eprosartan<br>Fosinopril<br>Irbesartan<br>Lisinopril<br>Losartan<br>Moexipril<br>Olmesartan<br>Perindopril<br>Quinapril<br>Ramipril<br>Telmisartan<br>Trandolapril<br>Valsartan |

**Appendix Table 2.** The ICD-10-CM, DRG, and ICD-10-PCS Procedure codes for pregnancy.

|  | ICD-10-CM | DRG | ICD-10-PCS<br>Procedure |
| --- | --- | --- | --- |
| Pregnancy | O00-O99, O9A1-O9A5, Z33, Z34, Z36, Z37, Z3201, Z322, Z39, F53, A34 | 765, 766, 767, 768, 769, 770, 771, 772, 773, 775, 776, 777, 779, 780, 781, 782 | 10A0, 10D00Z0, 10D00Z1, 10D00Z2, 10D07Z3, 10D07Z4, 10D07Z5, 10D07Z6, 10D07Z7, 10D07Z8, 10E0XZZ |

**Appendix Table 3.**
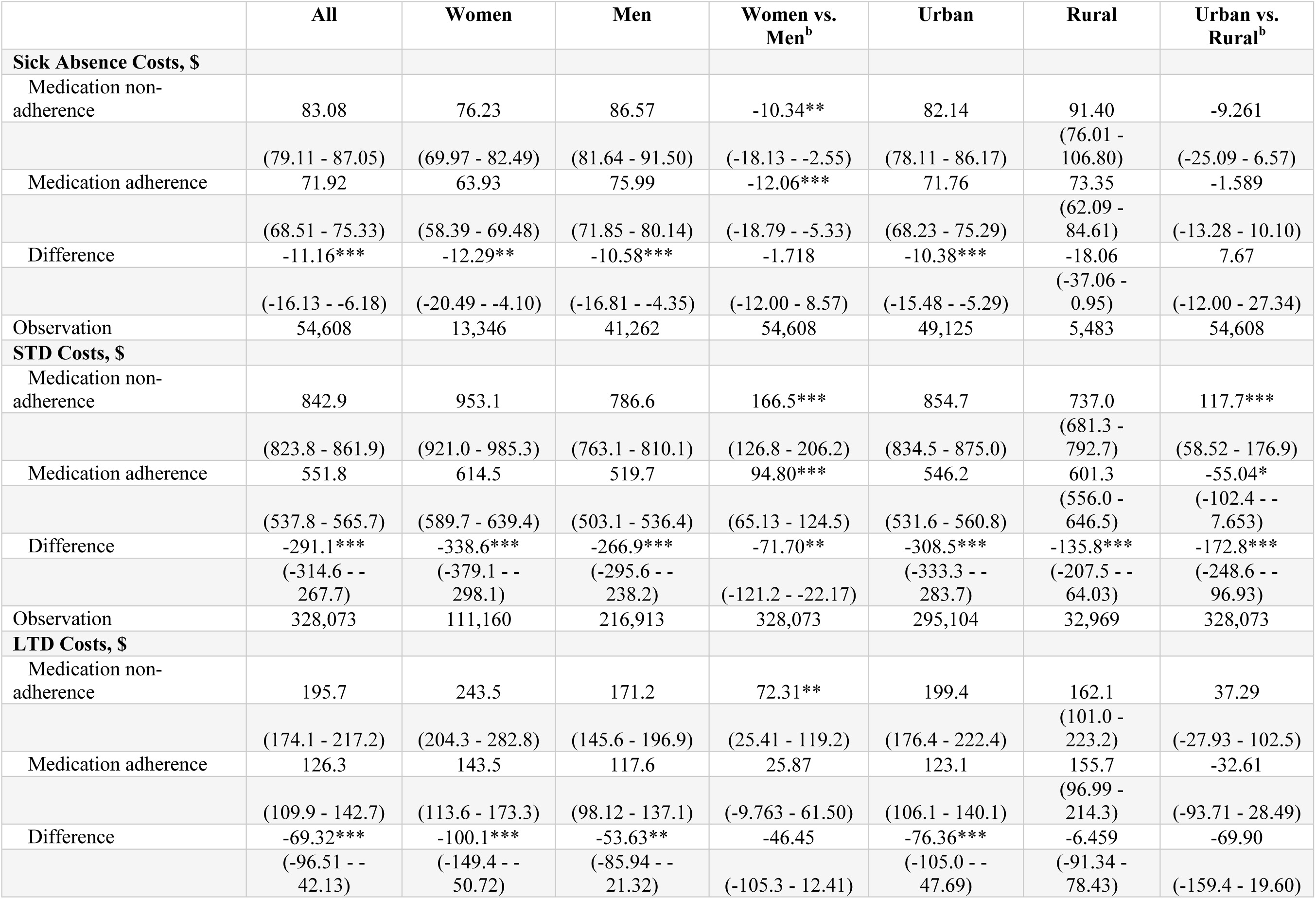
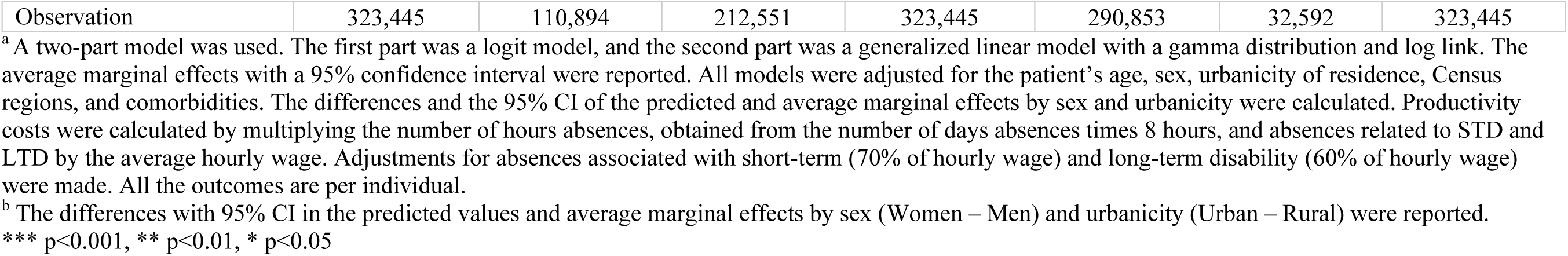
Productivity costs (per individual) associated with adherence to antihypertensives in 2019.^a^.

**Appendix Table 4.** Logistic regression estimates for all covariates used to calculate propensity scores^a^.

| **VARIABLES** | **Coefficient** | **Std.**<br>**err.** | **z** | **P>z** | **[95% conf.**<br>**interval]** |  |
| --- | --- | --- | --- | --- | --- | --- |
| Age 55–64 | 1.24 | 0.02 | 68.96 | <0.001 | 1.21 | 1.28 |
| Age 45–54 | 0.90 | 0.02 | 50.48 | <0.001 | 0.87 | 0.94 |
| Female | -0.27 | 0.03 | -9.25 | <0.001 | -0.32 | -0.21 |
| Age 35–44 | 0.49 | 0.02 | 27.07 | <0.001 | 0.45 | 0.52 |
| South | -0.32 | 0.02 | -13.78 | <0.001 | -0.37 | -0.28 |
| Congestive heart failure | -0.26 | 0.03 | -9.23 | <0.001 | -0.32 | -0.21 |
| Diabetes without chronic complications | 0.20 | 0.01 | 21.27 | <0.001 | 0.18 | 0.22 |
| Hemiplegia or paraplegia | -0.70 | 0.08 | -8.76 | <0.001 | -0.86 | -0.55 |
| Myocardial infarction | -0.16 | 0.03 | -4.67 | <0.001 | -0.22 | -0.09 |
| Chronic pulmonary disease | -0.22 | 0.02 | -10.10 | <0.001 | -0.27 | -0.18 |
| West | -0.16 | 0.02 | -9.21 | <0.001 | -0.19 | -0.13 |
| Cerebrovascular disease | -0.44 | 0.05 | -8.03 | <0.001 | -0.54 | -0.33 |
| Mild liver disease | -0.09 | 0.03 | -2.69 | 0.01 | -0.15 | -0.02 |
| Metastatic solid tumor | -0.37 | 0.06 | -6.18 | <0.001 | -0.48 | -0.25 |
| Moderate or severe liver disease | 0.31 | 0.36 | 0.87 | 0.39 | -0.39 | 1.01 |
| Midwest | -0.07 | 0.01 | -5.78 | <0.001 | -0.09 | -0.04 |
| Urban | -0.09 | 0.02 | -4.94 | <0.001 | -0.12 | -0.05 |
| Peptic ulcer | 0.35 | 0.25 | 1.36 | 0.17 | -0.15 | 0.84 |
| Renal disease | -0.12 | 0.04 | -3.52 | <0.001 | -0.19 | -0.05 |
| Any malignancy | 0.08 | 0.04 | 2.10 | 0.04 | 0.01 | 0.16 |
| Diabetes with chronic complications | 0.22 | 0.04 | 6.03 | <0.001 | 0.15 | 0.29 |
| HIV/AIDS | 0.22 | 0.07 | 3.23 | <0.001 | 0.09 | 0.36 |
| Auto Interaction: diabetes without chronic complications*diabetes with chronic complications | -0.38 | 0.04 | -9.18 | <0.001 | -0.46 | -0.30 |
| Auto Interaction: females*west region | 0.13 | 0.02 | 5.50 | <0.001 | 0.08 | 0.17 |
| Auto Interaction: females*age group 55 | 0.12 | 0.02 | 6.44 | <0.001 | 0.09 | 0.16 |
| Auto Interaction: females*any malignancy | 0.15 | 0.04 | 3.64 | <0.001 | 0.07 | 0.24 |
| Auto Interaction: south*urban | 0.09 | 0.02 | 4.23 | <0.001 | 0.05 | 0.14 |
| Auto Interaction: south*chronic pulmonary disease | 0.14 | 0.03 | 4.08 | <0.001 | 0.07 | 0.20 |
| Auto Interaction: female*south | -0.05 | 0.02 | -3.33 | <0.001 | -0.08 | -0.02 |
| Auto Interaction: congestive heart failure*renal disease | -0.22 | 0.07 | -3.26 | <0.001 | -0.36 | -0.09 |
| Auto Interaction: congestive heart failure*myocardial infarction | -0.22 | 0.07 | -3.35 | <0.001 | -0.35 | -0.09 |
| Auto Interaction: mild liver disease*any malignancy | -0.30 | 0.09 | -3.18 | <0.001 | -0.48 | -0.11 |
| Auto Interaction: west*diabetes with complications | 0.18 | 0.05 | 3.56 | <0.001 | 0.08 | 0.28 |
| Auto Interaction: age group 45*female | 0.06 | 0.02 | 2.99 | <0.001 | 0.02 | 0.10 |
| Auto Interaction: urban*peptic ulcer | -0.73 | 0.27 | -2.73 | 0.01 | -1.25 | -0.21 |
| Auto Interaction: age group 35*diabetes with complications | -0.18 | 0.06 | -3.26 | 0.00 | -0.29 | -0.07 |
| Auto Interaction: age group 55*west region | -0.06 | 0.02 | -2.83 | 0.01 | -0.10 | -0.02 |
| Auto Interaction: female*HIV/AIDS | -0.48 | 0.17 | -2.74 | 0.01 | -0.82 | -0.14 |
| Auto Interaction: mild liver disease*midwest region | -0.15 | 0.06 | -2.51 | 0.01 | -0.27 | -0.03 |
| Auto Interaction: moderate or severe liver disease*urban | -0.90 | 0.37 | -2.43 | 0.02 | -1.63 | -0.17 |
| Auto Interaction: south*renal disease | 0.10 | 0.04 | 2.39 | 0.02 | 0.02 | 0.18 |
| Auto Interaction: renal disease*any malignancy | -0.19 | 0.09 | -2.18 | 0.03 | -0.35 | -0.02 |
| Auto Interaction: age group 35*cerebrovascular disease | 0.39 | 0.17 | 2.28 | 0.02 | 0.05 | 0.72 |
| Auto Interaction: female*renal disease | 0.11 | 0.05 | 2.32 | 0.02 | 0.02 | 0.20 |
| Auto Interaction: diabetes without complications*myocardial infarction | -0.18 | 0.06 | -2.89 | <0.001 | -0.29 | -0.06 |
| Auto Interaction: myocardial infarction*diabetes with complications | 0.24 | 0.09 | 2.52 | 0.01 | 0.05 | 0.42 |
| Auto Interaction: age group 35* HIV/AIDS | -0.32 | 0.15 | -2.14 | 0.03 | -0.61 | -0.03 |
| Auto Interaction: age group 35*renal disease | 0.13 | 0.06 | 2.11 | 0.04 | 0.01 | 0.26 |
| Auto Interaction: female*myocardial infarction | -0.13 | 0.06 | -2.06 | 0.04 | -0.25 | -0.01 |
| Auto Interaction: age group 55*any malignancy | -0.09 | 0.04 | -2.03 | 0.04 | -0.17 | 0.00 |
| Auto Interaction: female*urban | -0.05 | 0.02 | -1.96 | 0.05 | -0.10 | 0.00 |
| Constant | -0.45 | 0.03 | -17.86 | <0.001 | -0.50 | -0.40 |

<sup>a</sup> This table shows logistic regression coefficients, standard error, z score, P-values, and confidence intervals for all covariates used in the final model obtained by applying iterative propensity score calculation method suggested by Imbens and Rubin, 2015.

**Appendix Table 5.** Propensity score estimated from logistic regression and calculated overlap weights for samples with medication adherence and medication non-adherence^a^.

|  | N | Q1 | Q2<br>(median) | Q3 | Inter quartile range (IQR) | Mean | SD | Min | Max |
| --- | --- | --- | --- | --- | --- | --- | --- | --- | --- |
| *Propensity score* |  |  |  |  |  |  |  |  |  |
| Medication non-adherence | 173189 | 0.457 | 0.535 | 0.611 | 0.154 | 0.522 | 0.105 | 0.079 | 0.757 |
| Medication adherence | 206310 | 0.518 | 0.575 | 0.620 | 0.102 | 0.562 | 0.090 | 0.117 | 0.758 |
| Total | 379499 | 0.472 | 0.559 | 0.618 | 0.146 | 0.544 | 0.099 | 0.079 | 0.758 |
| *Overlap weights* |  |  |  |  |  |  |  |  |  |
| Medication non-adherence | 173189 | 0.457 | 0.535 | 0.611 | 0.154 | 0.522 | 0.105 | 0.079 | 0.757 |
| Medication adherence | 206310 | 0.380 | 0.425 | 0.482 | 0.102 | 0.438 | 0.090 | 0.242 | 0.883 |
| Total | 379499 | 0.389 | 0.465 | 0.559 | 0.171 | 0.477 | 0.106 | 0.079 | 0.883 |

<sup>a</sup> This table shows the estimated propensity scores (logit) obtained from the logistic regression (shown in Appendix Table 4) and corresponding overlap weights for overall sample and for samples with medication adherence and non-adherence.

**Appendix Table 6.** Comparing unweighted and overlap-weighted samples to assess balance on covariates used in the main model^a^.

|  | Unweighted/Original Sample |  |  |  | Weighted Sample |  |  |  |
| --- | --- | --- | --- | --- | --- | --- | --- | --- |
|  | Medication adherence | Medication non-adherence | Standardized diff. | P value | Medication adherence | Medication non-adherence | Standardized diff. | P value |
| Age groups |  |  |  |  |  |  |  |  |
| 18-34 | 2.80% | 7.30% | -20.6% | <0.001 | 4.4% | 4.4% | 0.0% | 1 |
| 35-44 | 12.80% | 20.40% | -20.7% | <0.001 | 16.5% | 16.5% | 0.0% | 1 |
| 45-54 | 37.00% | 38.10% | -2.2% | <0.001 | 38.7% | 38.7% | 0.0% | 1 |
| 55-64 | 47.40% | 34.20% | 27.1% | <0.001 | 40.4% | 40.4% | 0.0% | 1 |
| Female | 31.40% | 36.70% | -11.3% | <0.001 | 34.0% | 34.0% | 0.0% | 1 |
| Urban Residency | 89.70% | 90.30% | -2.0% | <0.001 | 90.0% | 90.0% | 0.0% | 1 |
| Northeast | 15.10% | 12.40% | 7.6% | <0.001 | 13.7% | 13.6% | 0.3% | 0.311 |
| Midwest | 26.40% | 24.10% | 5.3% | <0.001 | 25.2% | 25.2% | 0.0% | 1 |
| South | 45.60% | 50.90% | -10.6% | <0.001 | 48.3% | 48.3% | 0.0% | 1 |
| West | 12.70% | 12.30% | 1.3% | <0.001 | 12.6% | 12.6% | 0.0% | 1 |
| Comorbidities |  |  |  |  |  |  |  |  |
| Myocardial infarction | 2.0% | 2.0% | -2.5% | <0.001 | 2.0% | 2.0% | 0.0% | 1 |
| Congestive heart failure | 2.0% | 2.0% | -4.2% | <0.001 | 2.0% | 2.0% | 0.0% | 1 |
| Peripheral vascular disease | 1.0% | 1.0% | -0.1% | 0.866 | 1.0% | 1.0% | -0.1% | 0.66 |
| Cerebrovascular disease | 0.0% | 1.0% | -3.3% | <0.001 | 0.0% | 0.0% | 0.0% | 1 |
| Dementia | 0.0% | 0.0% | -0.7% | 0.028 | 0.0% | 0.0% | -0.8% | 0.016 |
| Chronic pulmonary disease | 4.0% | 5.0% | -3.6% | <0.001 | 4.0% | 4.0% | 0.0% | 1 |
| Rheumatic disease | 1.0% | 1.0% | -0.5% | 0.123 | 1.0% | 1.0% | 0.0% | 0.932 |
| Peptic ulcer disease | 0.0% | 0.0% | -1.6% | <0.001 | 0.0% | 0.0% | 0.0% | 1 |
| Mild liver disease | 1.0% | 2.0% | -2.4% | <0.001 | 2.0% | 2.0% | 0.0% | 1 |
| Diabetes without chronic complication | 20.0% | 17.0% | 8.4% | <0.001 | 18.0% | 18.0% | 0.0% | 1 |
| Diabetes with chronic complication | 4.0% | 4.0% | 2.1% | <0.001 | 4.0% | 4.0% | 0.0% | 1 |
| Hemiplegia or paraplegia | 0.0% | 0.0% | -3.8% | <0.001 | 0.0% | 0.0% | 0.0% | 1 |
| Renal disease | 3.0% | 3.0% | -0.9% | 0.009 | 3.0% | 3.0% | 0.0% | 1 |
| Any malignancy | 3.0% | 3.0% | 2.2% | <0.001 | 3.0% | 3.0% | 0.0% | 1 |
| Moderate or severe liver disease | 0.0% | 0.0% | -2.2% | <0.001 | 0.0% | 0.0% | 0.0% | 1 |
| Metastatic solid tumor | 0.0% | 0.0% | -1.6% | <0.001 | 0.0% | 0.0% | 0.0% | 1 |
| HIV/AIDS | 0.0% | 0.0% | 0.2% | 0.525 | 0.0% | 0.0% | 0.0% | 1 |

<sup>a</sup> This table shows standardized difference for means in unweighted (original) and overlap-weighted samples to assess balance between population with and without medication adherence. The associated standardized difference and p-values for overlap-weighted samples suggest that the sample is balanced on all covariates used in the model and indicate validity of overlap-weighting model to adjust for measured confounders.

**Appendix Table 7.** Underlying medication adherence rates by sex and urbanicity (Unadjusted)^a^.

|  | **Male** | **Female** | ***P-values*** |
| --- | --- | --- | --- |
|  | N=251,258 | N=128,245 |  |
| **Medication adherence<sup>b</sup>, N (%)** | 141,631 (56.37%) | 64,679 (50.43%) | <0.001 |
|  | **Rural** | **Urban** |  |
|  | N=38,211 | N=341,292 |  |
| **Medication adherence, N (%)** | 21,332 (55.83%) | 184,978 (54.20%) | <0.001 |

<sup>a</sup> The Pearson's Chi-square test was used to test differences in proportions for medication adherence by sex (male vs. female) and urbanicity (rural vs. urban) of residence.
<sup>b</sup> Adherence to antihypertensives if the average medication possession ratio (MPR) of the 7 Therapeutic Classes was greater than equal to 80%; otherwise, medication non-adherence.

**Appendix Table 8.** Association of individuals’ demographics and comorbidities with medication adherence (Adjusted)**^a^**.

| Models | OLS | Logit | Probit |
| --- | --- | --- | --- |
| Dependent variable | Medication adherence<sup>b</sup> | Medication adherence | Medication adherence |
| Sex |  |  |  |
| Male (reference) |  |  |  |
| **Female** | **-0.0583\*\*\*** | **-0.0583\*\*\*** | **-0.0583\*\*\*** |
|  | **(-0.0616 - - 0.0550)** | **(-0.0616 - - 0.0550)** | **(-0.0616 - - 0.0550)** |
| Urbanicity of residence |  |  |  |
| Rural (reference) |  |  |  |
| **Urban** | **-0.0138\*\*\*** | **-0.0138\*\*\*** | **-0.0138\*\*\*** |
|  | **(-0.0189 - - 0.00856)** | **(-0.0189 - - 0.00857)** | **(-0.0190 - - 0.00858)** |
| Age groups |  |  |  |
| Aged 18-34 (reference) |  |  |  |
| Aged 35-44 | 0.112*** | 0.116*** | 0.115*** |
|  | (0.104 - 0.120) | (0.107 - 0.124) | (0.107 - 0.123) |
| Aged 45-54 | 0.219*** | 0.220*** | 0.220*** |
|  | (0.212 - 0.227) | (0.212 - 0.227) | (0.212 - 0.227) |
| Aged 55-64 | 0.305*** | 0.305*** | 0.305*** |
|  | (0.298 - 0.313) | (0.297 - 0.313) | (0.298 - 0.313) |
| Regions |  |  |  |
| Northeast (Reference) |  |  |  |
| Midwest | -0.0154*** | -0.0155*** | -0.0155*** |
|  | (-0.0206 - - 0.0102) | (-0.0208 - - 0.0103) | (-0.0207 - - 0.0103) |
| South | -0.0584*** | -0.0585*** | -0.0585*** |
|  | (-0.0632 - - 0.0537) | (-0.0632 - - 0.0537) | (-0.0632 - - 0.0537) |
| West | -0.0310*** | -0.0312*** | -0.0312*** |
|  | (-0.0370 - - | (-0.0372 - - | (-0.0373 - - |

|  | 0.0249) | 0.0251) | 0.0252) |
| --- | --- | --- | --- |
| Comorbidities |  |  |  |
| Myocardial infarction | -0.0619*** | -0.0615*** | -0.0616*** |
|  | (-0.0738 - -<br>0.0500) | (-0.0733 - -<br>0.0497) | (-0.0734 - -<br>0.0497) |
| Congestive heart failure | -0.0802*** | -0.0799*** | -0.0799*** |
|  | (-0.0919 - -<br>0.0686) | (-0.0915 - -<br>0.0682) | (-0.0916 - -<br>0.0683) |
| Peripheral vascular disease | -0.00574 | -0.00566 | -0.00571 |
|  | (-0.0191 -<br>0.00765) | (-0.0191 -<br>0.00775) | (-0.0191 -<br>0.00768) |
| Cerebrovascular disease | -0.0969*** | -0.0973*** | -0.0967*** |
|  | (-0.121 - -<br>0.0727) | (-0.122 - -<br>0.0729) | (-0.121 - -<br>0.0725) |
| Dementia | -0.171* | -0.170* | -0.172* |
|  | (-0.308 - -<br>0.0339) | (-0.310 - -<br>0.0312) | (-0.310 - -<br>0.0325) |
| Chronic pulmonary disease | -0.0393*** | -0.0391*** | -0.0392*** |
|  | (-0.0470 - -<br>0.0315) | (-0.0468 - -<br>0.0314) | (-0.0469 - -<br>0.0315) |
| Rheumatic disease | 0.00177 | 0.00167 | 0.00164 |
|  | (-0.0135 -<br>0.0171) | (-0.0136 -<br>0.0170) | (-0.0137 -<br>0.0169) |
| Peptic ulcer disease | -0.0762*** | -0.0759*** | -0.0759*** |
|  | (-0.112 - -<br>0.0404) | (-0.112 - -<br>0.0400) | (-0.112 - -<br>0.0400) |
| Mild liver disease | -0.0360*** | -0.0358*** | -0.0360*** |
|  | (-0.0487 - -<br>0.0234) | (-0.0485 - -<br>0.0232) | (-0.0486 - -<br>0.0233) |
| Diabetes without chronic complication | 0.0422*** | 0.0424*** | 0.0424*** |
|  | (0.0380 - 0.0464) | (0.0382 - 0.0467) | (0.0381 - 0.0467) |
| Diabetes with chronic complication | -0.0110* | -0.0110* | -0.0110* |
|  | (-0.0195 - -<br>0.00240) | (-0.0196 - -<br>0.00239) | (-0.0196 - -<br>0.00241) |
| Hemiplegia or paraplegia | -0.162*** | -0.167*** | -0.166*** |
|  | (-0.198 - -0.126) | (-0.205 - -0.130) | (-0.203 - -0.129) |
| Renal disease | -0.0148** | -0.0149** | -0.0149** |
|  | (-0.0249 - -<br>0.00467) | (-0.0250 - -<br>0.00480) | (-0.0250 - -<br>0.00480) |
| Any malignancy | 0.0161** | 0.0163** | 0.0161** |
|  | (0.00591 -<br>0.0263) | (0.00604 -<br>0.0265) | (0.00589 -<br>0.0263) |
| Moderate or severe liver disease | -0.128*** | -0.129*** | -0.130*** |
|  | (-0.174 - -<br>0.0824) | (-0.176 - -<br>0.0828) | (-0.176 - -<br>0.0835) |
| Metastatic solid tumor | -0.0949*** | -0.0941*** | -0.0940*** |
|  | (-0.122 - -<br>0.0674) | (-0.122 - -<br>0.0667) | (-0.121 - -<br>0.0667) |
| HIV/AIDS | 0.0219 | 0.0219 | 0.0219 |
|  | (-0.00516 -<br>0.0490) | (-0.00524 -<br>0.0491) | (-0.00530 -<br>0.0492) |
| Observations | 379,503 | 379,503 | 379,503 |

<sup>a</sup> This table presents the association of individuals' demographics and comorbidities with medication adherence. We provided the average marginal effects with 95% confidence intervals from linear regression, logit, and probit models.
<sup>b</sup> Adherence to antihypertensives if the average medication possession ratio (MPR) of the 7 Therapeutic Classes was greater than equal to 80%; otherwise, medication non-adherence.
\*\*\* $P < 0.001$ , \*\* $P < 0.01$ , \* $P < 0.05$

**Appendix Table 9.** Sensitivity analysis using propensity score based overlap-weighting: total medical costs (per individual) associated with adherence to antihypertensives in 2019^a^.

|  | All | Women | Men | Urban | Rural |
| --- | --- | --- | --- | --- | --- |
| **Total Medical Costs, \$** | | | | | |
| Medication non-adherence | 18,696 | 19,795 | 18,131 | 18,933 | 16,562 |
|  | (18,290 - 19,103) | (19,330 - 20,259) | (17,692 - 18,570) | (18,509 - 19,358) | (16,000 - 17,125) |
| Medication adherence | 17,318 | 18,666 | 16,625 | 17,508 | 15,612 |
|  | (16,938 - 17,698) | (18,229 - 19,103) | (16,220 - 17,029) | (17,112 - 17,903) | (15,117 - 16,108) |
| Difference | -1,378*** | -1,129*** | -1,506*** | -1,426*** | -949.7** |
|  | (-1,628 - -1,128) | (-1,543 - -714.5) | (-1,818 - -1,194) | (-1,693 - -1,158) | (-1,618 - -281.8) |
| Observation | 379,503 | 128,245 | 251,258 | 341,292 | 38,211 |

<sup>a</sup> We applied the overlap weighting method, calculating weights derived from logistic regression, with adherence to antihypertensives as the outcome. A generalized linear model with a gamma distribution and log link was used. The average marginal effects with 95% confidence intervals (CI) were reported. All models were adjusted for the patient's age, sex, urbanicity of residence, Census regions, and comorbidities. The differences and the 95% CI of the predicted and average marginal effects by sex and urbanicity were calculated. All the outcomes are per individual.

**Appendix Table 10.** Sensitivity analysis using propensity score based overlap-weighting model: health care utilization (per 1,000 individuals) associated with adherence to antihypertensives in 2019^a^.

|  | All | Women | Men | Urban | Rural |
| --- | --- | --- | --- | --- | --- |
| **Emergency Department Visits** |  |  |  |  |  |
| Medication non-adherence | 433 | 539 | 379 | 434 | 431 |
|  | (428 - 439) | (529 - 549) | (373 - 385) | (428 - 439) | (416 - 446) |
| Medication adherence | 264 | 316 | 238 | 263 | 273 |
|  | (261 - 268) | (309 - 323) | (234 - 242) | (260 - 267) | (263 - 283) |
| Difference | -169*** | -223*** | -141*** | -170*** | -158*** |
|  | (-175 - -163) | (-234 - -211) | (-148 - -134) | (-177 - -164) | (-176 - -139) |
| Observation | 379,503 | 128,245 | 251,258 | 341,292 | 38,211 |
| **Inpatient Admissions** |  |  |  |  |  |
| Medication non-adherence | 190 | 160 | 206 | 195 | 147 |
|  | (182 - 199) | (153 - 167) | (195 - 216) | (186 - 204) | (137 - 156) |
| Medication adherence | 108 | 97 | 114 | 110 | 88 |
|  | (103 - 113) | (92 - 102) | (108 - 120) | (105 - 116) | (82 - 94) |
| Difference | -82*** | -63*** | -92*** | -85*** | -59*** |
|  | (-87 - -77) | (-69 - -57) | (-99 - -85) | (-90 - -79) | (-69 - -49) |
| Observation | 379,503 | 128,245 | 251,258 | 341,292 | 38,211 |

Abbreviations: ED, emergency department.
<sup>a</sup> We applied the overlap weighting method, calculating weights derived from logistic regression, with adherence to antihypertensives as the outcome. A negative binomial regression was used. All models were adjusted for the patient's age, sex, urbanicity of residence, Census regions, and comorbidities. We reported the average marginal effects with a 95% confidence interval. The differences and the 95% CI of the predicted and average marginal effects by sex and urbanicity were calculated. All the outcomes are per 1,000 individuals.

**Appendix Table 11.** Sensitivity analysis using propensity score based overlap-weighting model: productivity losses (per 1,000 individuals) associated with adherence to antihypertensives in 2019^a^.

|  | All | Women | Men | Urban | Rural |
| --- | --- | --- | --- | --- | --- |
| **Number of Sick Absences** |  |  |  |  |  |
| Medication non-adherence | 308 | 327 | 299 | 300 | 387 |
|  | (299 - 317) | (314 - 340) | (289 - 308) | (291 - 308) | (353 - 420) |
| Medication adherence | 289 | 305 | 281 | 285 | 329 |
|  | (281 - 297) | (293 - 318) | (272 - 289) | (276 - 293) | (303 - 354) |
| Difference | -19*** | -22** | -18*** | -15*** | -58** |
|  | (-28 - -11) | (-38 - -6) | (-28 - -8) | (-23 - -7) | (-98 - -19) |
| Observation | 54,608 | 13,346 | 41,262 | 49,125 | 5,483 |
| **Number of STD** |  |  |  |  |  |
| Medication non-adherence | 4,530 | 4,335 | 4,631 | 4,563 | 4,237 |
|  | (4,431 - 4,629) | (4,212 - 4,457) | (4,509 - 4,752) | (4,459 - 4,666) | (3,975 - 4,499) |
| Medication adherence | 4,087 | 3,944 | 4,160 | 4,098 | 3,985 |
|  | (3,994 - 4,180) | (3,823 - 4,066) | (4,048 - 4,273) | (4,002 - 4,195) | (3,741 - 4,228) |
| Difference | -443*** | -390*** | -470*** | -464*** | -252 |
|  | (-550 - -336) | (-0,542 - -0,239) | (-612 - -328) | (-577 - -351) | (-603 - 99) |
| Observation | 328,073 | 111,160 | 216,913 | 295,104 | 32,969 |
| **Number of LTD** |  |  |  |  |  |
| Medication non-adherence | 1,519 | 1,239 | 1,664 | 1,497 | 1,722 |
|  | (1,343 - 1,696) | (1,043 - 1,434) | (1,443 - 1,884) | (1,317 - 1,677) | (1,304 - 2,140) |
| Medication adherence | 1,637 | 1,481 | 1,716 | 1,603 | 1,939 |
|  | (1,438 - 1,835) | (1,228 - 1,734) | (1,476 - 1,957) | (1,400 - 1,806) | (1,428 - 2,450) |
| Difference | 117 | 243 | 53 | 106 | 217 |
|  | (-85 - 319) | (-38 - 523) | (-216 - 321) | (-107 - 319) | (-404 - 838) |
| Observation | 323,445 | 110,894 | 212,551 | 290,853 | 32,592 |

Abbreviations: LTD, long-term disability; STD, short-term disability.
<sup>a</sup> We applied the overlap weighting method, calculating weights derived from logistic regression, with adherence to antihypertensives as the outcome variable. An exponential hurdle model was used. All models were adjusted for the patient's age, sex, urbanicity of residence, Census regions, and comorbidities. We reported the average marginal effects with a 95% confidence interval. The differences and the 95% CI of the predicted and average marginal effects by sex and urbanicity were calculated. All the outcomes are per 1,000 individuals.

**Appendix Table 12.** Sensitivity analysis using propensity score based overlap-weighting: productivity costs (per individual) associated with adherence to antihypertensives in 2019.^a^.

|  | All | Women | Men | Urban | Rural |
| --- | --- | --- | --- | --- | --- |
| **Sick Absence Costs, \$** | | | | | |
| Medication non-adherence | 84.58 | 73.52 | 90.22 | 83.12 | 97.64 |
|  | (80.50 - 88.65) | (67.55 - 79.48) | (85.21 - 95.24) | (79.04 - 87.19) | (81.85 - 113.4) |
| Medication adherence | 70.97 | 59.67 | 76.74 | 69.70 | 82.35 |
|  | (67.78 - 74.16) | (55.23 - 64.11) | (72.64 - 80.83) | (66.49 - 72.90) | (70.21 - 94.50) |
| Difference | -13.61*** | -13.85*** | -13.49*** | -13.42*** | -15.28 |
|  | (-18.35 - -8.871) | (-21.14 - -6.557) | (-19.57 - -7.408) | (-18.21 - -8.630) | (-35.00 - 4.433) |
| Observation | 54,608 | 13,346 | 41,262 | 49,125 | 5,483 |
| **STD Costs, \$** | | | | | |
| Medication non-adherence | 839.3 | 940.9 | 787.4 | 848.6 | 755.7 |
|  | (819.9 - 858.6) | (909.1 - 972.8) | (763.4 - 811.4) | (828.2 - 869.1) | (695.9 - 815.4) |
| Medication adherence | 547.5 | 619.4 | 510.8 | 543.8 | 580.8 |
|  | (533.6 - 561.4) | (595.1 - 643.7) | (494.0 - 527.6) | (529.2 - 558.4) | (536.0 - 625.6) |
| Difference | -291.8*** | -321.5*** | -276.6*** | -304.8*** | -174.8*** |
|  | (-315.4 - -268.1) | (-361.4 - -281.6) | (-305.8 - -247.4) | (-329.7 - -279.9) | (-249.5 - -100.1) |
| Observation | 328,073 | 111,160 | 216,913 | 295,104 | 32,969 |
| **LTD Costs, \$** | | | | | |
| Medication non-adherence | 195.6 | 238.3 | 173.8 | 199.8 | 157.4 |
|  | (173.8 - 217.4) | (193.9 - 282.6) | (150.1 - 197.4) | (176.2 - 223.5) | (117.1 - 197.6) |
| Medication adherence | 126.2 | 142.0 | 118.1 | 122.8 | 156.3 |
|  | (113.4 - 139.0) | (121.1 - 163.0) | (101.8 - 134.4) | (109.2 - 136.4) | (117.2 - 195.4) |
| Difference | -69.39*** | -96.22*** | -55.69*** | -77.04*** | -1.056 |
|  | (-94.27 - -44.51) | (-145.2 - -47.28) | (-83.98 - -27.40) | (-103.9 - -50.18) | (-57.10 - 54.98) |
| Observation | 323,445 | 110,894 | 212,551 | 290,853 | 32,592 |

<sup>a</sup> We applied the overlap weighting method, calculating weights derived from logistic regression, with adherence to antihypertensives as the outcome variable. A two-part model was used. The first part was a logit model, and the second part was a generalized linear model with a gamma distribution and log link. The average marginal effects with a 95% confidence interval were reported. All models were adjusted for the patient's age, sex, urbanicity of residence, Census regions, and comorbidities. The differences and the 95% CI of the predicted and average marginal effects by sex and urbanicity were calculated. Productivity costs were calculated by multiplying the number of hours absences, obtained from the number of days absences times 8 hours, and absences related to STD and LTD by the average hourly wage. Adjustments for absences associated with short-term (70% of hourly wage) and long-term disability (60% of hourly wage) were made. All the outcomes are per individual.

**Appendix Figure 1:**
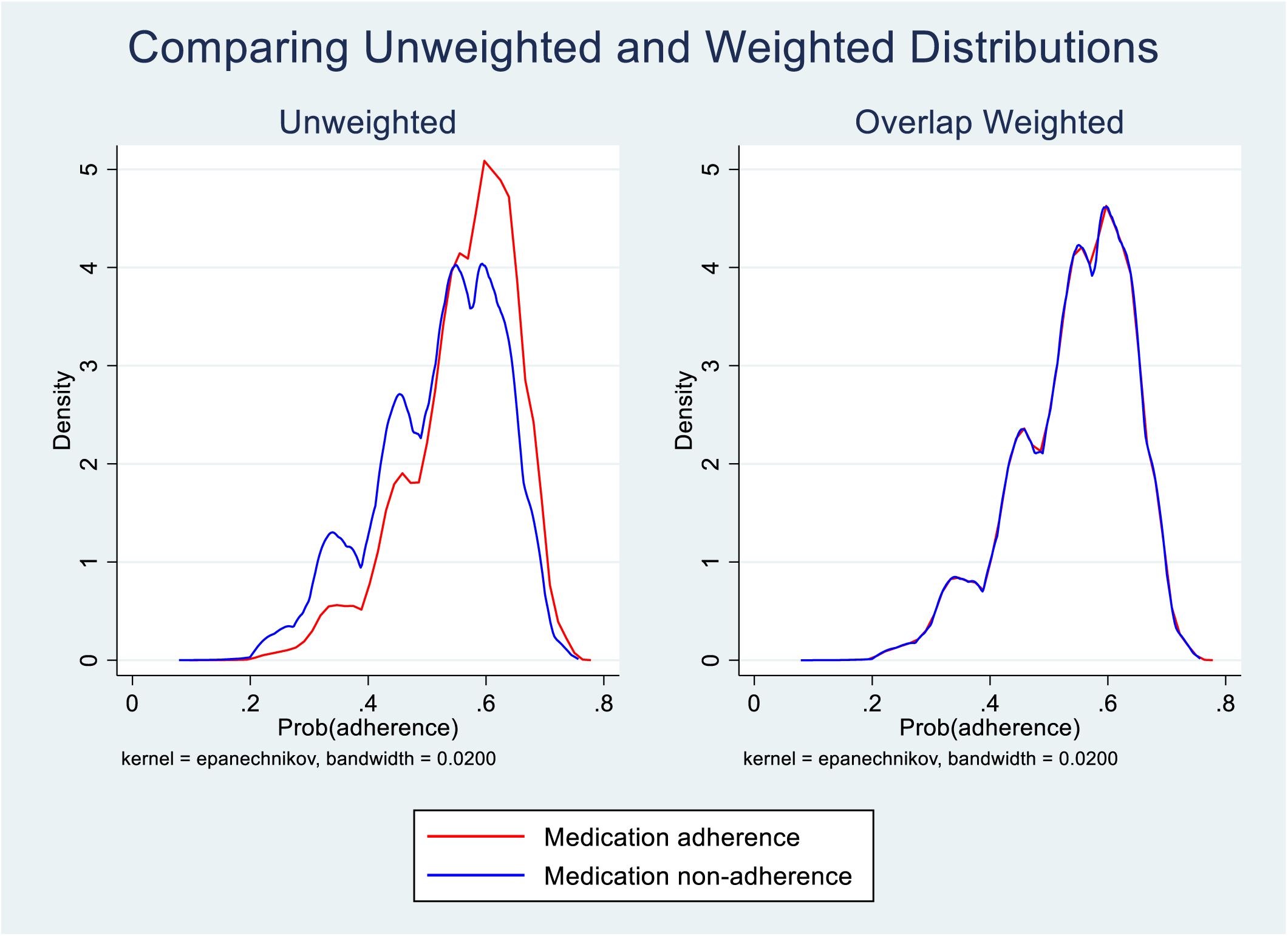
Difference in distribution of probability of adherence estimated from logistic regression in unweighted (original) and overlap-weighted samples^a^ ^a^ This figure shows the difference in probability distribution for unweighted and weighted samples among individuals with and without medication adherence. The distributions for overlap-weighted samples of the population with and without medication adherence suggest the validity of the overlap-weighting model for adjusting for measured confounders.

**Appendix Figure 2:**
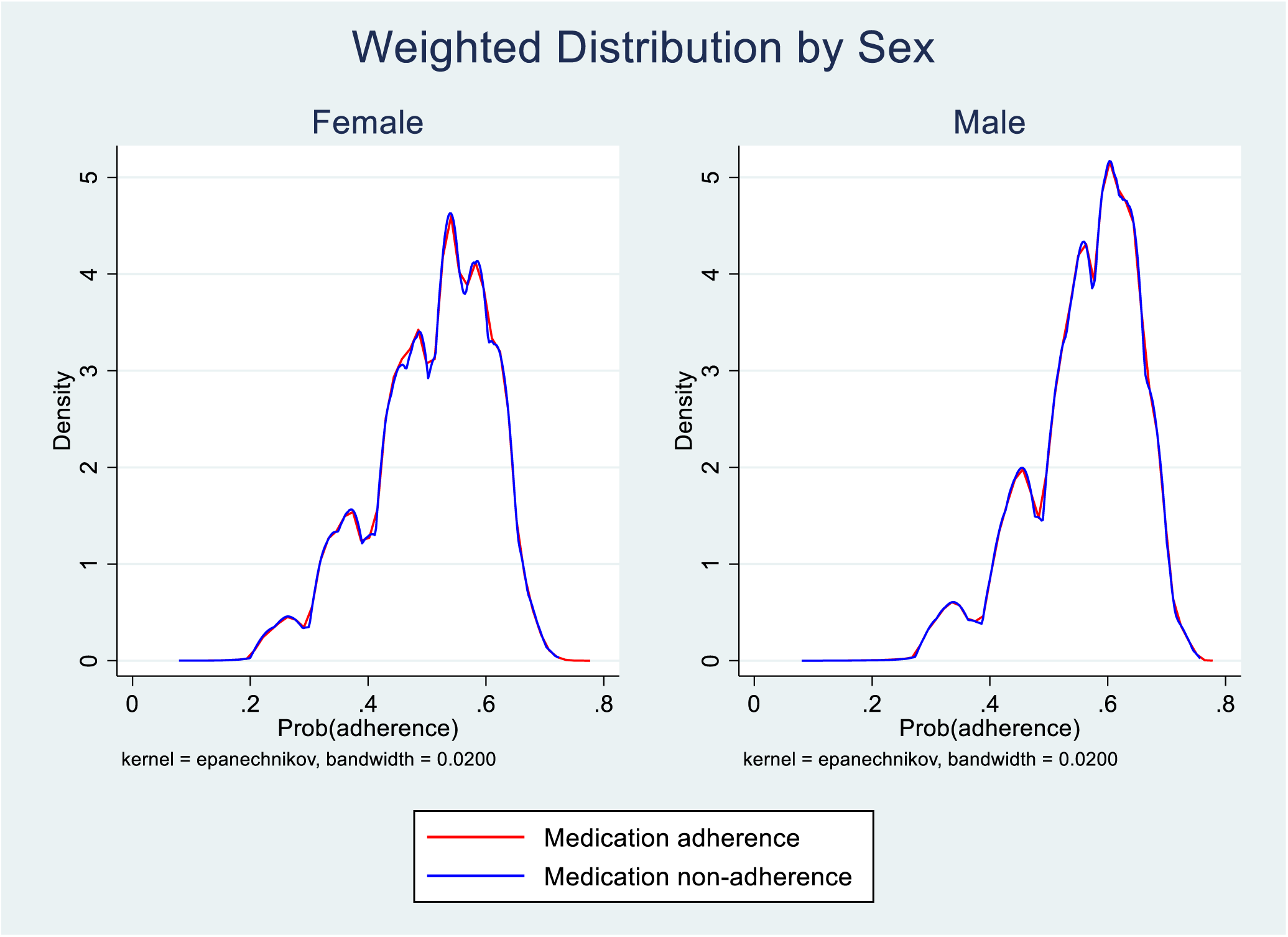
Difference in distribution of probability of adherence estimated from logistic regression in the overlap-weighted sample by sex^a^ ^a^ This figure shows the difference in probability distribution for overlap-weighted samples by sex among individuals with and without medication adherence. The distributions for overlap-weighted samples of the population with and without medication adherence suggest the validity of the overlap-weighting model for adjusting for measured confounders.

**Appendix Figure 3:**
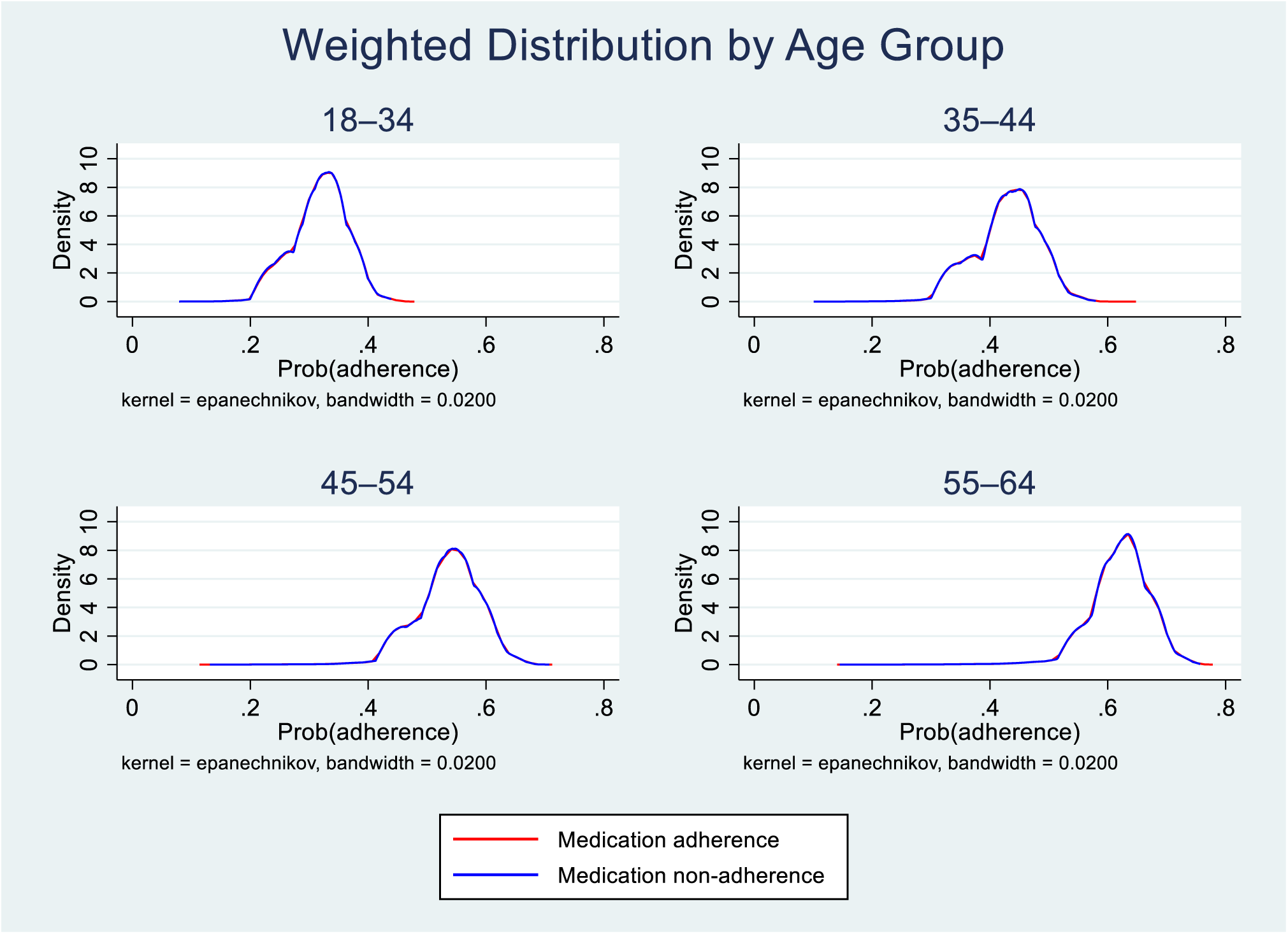
Difference in distribution of probability of adherence estimated from logistic regression in the overlap-weighted sample by age group^a^ ^a^ This figure shows the difference in probability distribution for overlap-weighted samples by age group among individuals with and without medication adherence. The distributions for overlap-weighted samples of the population with and without medication adherence suggest the validity of the overlap-weighting model for adjusting for measured confounders.

**Appendix Figure 4:**
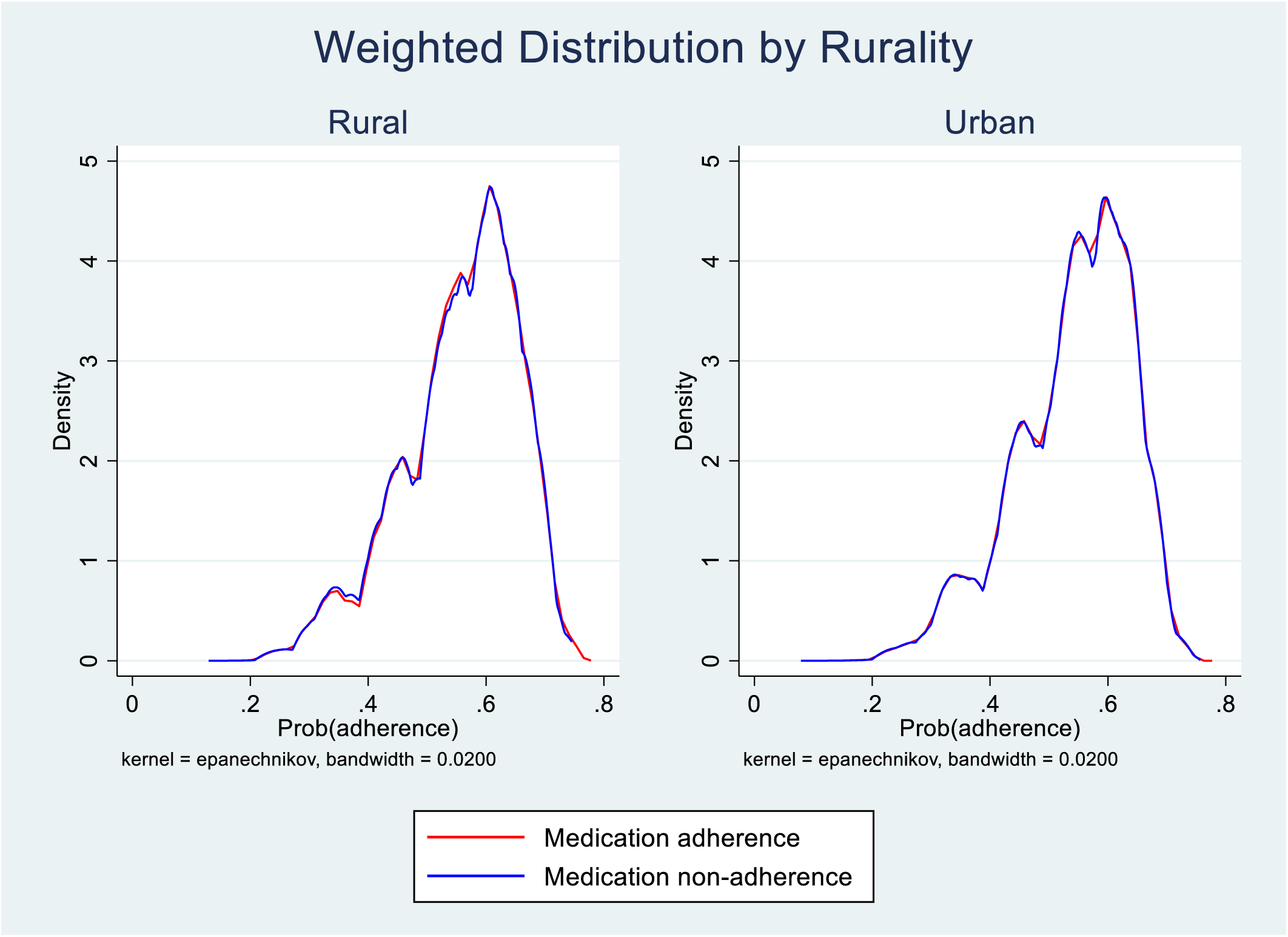
Difference in distribution of probability of adherence estimated from logistic regression in e -weighted sample by rurality of residence^a^ ^a^ This figure shows the difference in probability distribution for overlap-weighted samples by rurality among those with and without medication adherence. The distributions for overlap-weighted samples of population with and without medication adherence suggest validity of overlap-weighting model to adjust for measured confounders.

## Notes

### Competing Interest Statement

The authors have declared no competing interest.

### Clinical Trial

NA

### Funding Statement

The authors received no financial support for the research, authorship, and/or publication of this article.

### Author Declarations

This study involved secondary data analysis using de-identified information and was categorized as non-research and thus exempt from Institutional Review Board review.

